# Assessing Occupational Hazards and Factors Contributing to the Health Effects Faced By Female Market Traders in Nakasero Market, Central Kampala Uganda

**DOI:** 10.1101/2023.10.31.23297787

**Authors:** Waneloba Ronald

## Abstract

**Background:** In Uganda, the informal sector, particularly public markets, presents a concerning array of unregulated workplace hazards, predominantly affecting female traders. These hazards pose significant threats to the health of female market traders, who make up the majority of the workforce. However, there has been limited attention given to identifying and addressing the occupational risks faced by these women. This study aimed to assess the occupational hazards and the contributing factors to health effects among female market traders in Nakasero market, central Kampala.

**Methods:** A descriptive cross-sectional study was conducted among female market traders in Nakasero market, central Kampala. The study employed a mixed-methods approach, utilizing both quantitative and qualitative methods. Data collection involved face-to-face, interviewer-administered, semi-structured questionnaires and key informant interviews. Quantitative data were analyzed using EPIDATA 3.02, STATA 14, and Microsoft Excel, while qualitative data were subjected to deductive thematic analysis.

**Results:** The hazards encompassed biological risks, where a significant proportion reported suffering from mosquito bites within the market (87.36%), and urinary tract infections (UTIs) attributed to microorganisms stemming from unclean toilet facilities (80.46%). Furthermore, the investigation revealed the presence of physical, ergonomic, and psychological hazards, all of which posed a threat to the health of female market traders.

Various factors contributed to the health effects experienced by these traders. Notably, many of them endured prolonged working hours without adequate rest or sleep, with 47% of female market traders spending over 12 hours in the market. The study also found that a majority of respondents failed to use personal protective equipment (PPE), totaling 70.49%. Additionally, a substantial portion (42%) admitted to not practicing proper hand hygiene while in the market. Issues related to sanitary facilities were prevalent, including a shortage of toilet stalls, unclean conditions, and occasional water shortages. Poor market infrastructure, inadequate working space, and overcrowding further compounded the challenges faced by female market traders.

The health effects and problems reported by these traders encompassed a spectrum of issues, including musculoskeletal pain, malaria, UTIs, respiratory problems, COVID-19, skin conditions, headaches, and obesity.

**Conclusion:** This study underscores the ongoing occupational hazards and health effects faced by female market traders in public markets throughout Uganda, arising from a combination of various contributing factors. Addressing these issues is crucial to safeguard the well-being of these women and improve their working conditions.

## INTRODUCTION

Female participation in both rural and urban public markets has been increasing in the past few decades as population grows high in Uganda and outside world(Flaspöler et al. 2013).A recent estimate shows that the share of informal economy such as market areas and roadside vendors in sub-Saharan Africa is high up to 72% characterised by flexibility, job insecurity, unhealthy and unsafe settings(Uko, Akpanoyoro, and Ekpe 2020).For instance women are indeed more exposed to informal employment in more than 90% of sub-Saharan African countries, 89% of countries from south Asia and almost 75% of Latin American countries (Alfers 2009). By some estimates, 92 per cent of workers in sub-Saharan Africa are employed in the informal economy (Bonnet, Vanek, and Chen 2019).Uganda is considered the only African country on the top 10 list of countries with highest level of informal economic activity and the top with 94% of its population working in unregulated and untaxed jobs (worldatlas 2019).

Female traders in informal markets in developing countries is dominated by small firms/self-employment, low paid wage earnings and work extended hours, absence of regulatory coverage in terms of occupational safety and health, collective bargaining, and workers compensation and this makes workers situation especially for children and women more dire other nonstandard workers(Bonnet, Vanek, and Chen 2019).Female market traders contribute a lot to consumer economy and living standards of a large number of families that have poor education backgrounds and unemployed populations yet they are more prone sedentariness, at risk of obesity and associated to non-communicable chronic diseases, long working hours with physical inactivity, psychological stress due to low wages, unsafe work conditions and lack of job security(Amoako 2019).

Therefore this research focused on identifying the key factors that exposes female workers to occupational health hazards in the markets and this can help to determine the burden of preventable diseases, injuries and disabilities and can provide valuable new insights on health related quality in such occupations.

## BACKGROUND

Women’s occupational health has become an issue of public health concern in Uganda and at a global perspective. The international labour organization reported that globally more than 70% of market and street traders are women(Chen 2001). occupational health and safety of these women is at stake due to their increased female involvement in various productive activities, work that is unprotected, heavy workload and badly paid informal economy like market and street side trade(Amoako 2019).

A study done in Accra and Takoradi, Ghana shown that female market traders who works in these informal market areas and roadsides are at higher risk of environmental diseases, traffic accidents, fire hazards, crime and assault, weather related discomfort, and musculoskeletal injuries(Alfers 2009).

Another study on both men and women informal sector workers in Dar es Salaam, Tanzania revealed that workers in the informal sector are exposed to biological, mechanical, ergonomic, physical and psycho-social hazards against which they are poorly protected(Rajen N Naidoo 2009)

Locally, during Covid-19 pandemic in Uganda, unlike street vendors, women in the KAMPALA markets were permitted to work during the lockdown, on condition that they stayed in the market. However, female market traders decried the deplorable and unsafe state in which they worked. This was attributed by risk factors like the poor market structures rendering them vulnerable to health hazards that included; poor weather conditions, no bathrooms available so women have to bathe in the toilets and long working hours of day in day out, women suffered losses from damaged goods due to the reduction in the numbers of customers and the rising use of middle men who purchased directly from farmers(akinamamawafrica 2020).

The informal sector that includes street and market trade remains a critical source of livelihood for women, households, and the economy in Uganda(UBOS 2020). The sector contributes 54 percent of gross domestic product (GDP) and absorbs 87 percent of women workers(UBOS 2020). Over 50% of Uganda’s GDP is attributed to the informal sector and more than 80% of the labour force work is in the informal sector according to the Uganda Bureau of Statistics (<u>UBOS2014</u>). A report by the ILO stated that women-owned businesses by 2014 had outpaced male-owned businesses by 1.5 times (236% compared to 153 %.). According to the report, though women own 44% of the businesses, they are mainly engaged in self-employment (86.2% of working women)(Fred, Nduhura, and Ronald).

Various informal sector workplaces in Uganda are described, including home informal enterprises, public markets, displaying a wide range of poorly controlled work hazards, particularly welfare and hygiene, ergonomic and chemical hazards, worsened by poor work organization, and poor community environments and social infrastructures(Loewenson 2002; Kevin 2019).Marketers work in crowded conditions, street hawkers have very little access to hand-washing facilities. As a result, informal workers are among the most vulnerable to COVID-19’s health and economic shocks(Megersa 2020).

Nothing much is known about the occupational health of female market traders in informal sector more so the public markets. This research will seek to identify some of the major hazards and factors contributing to the health problems faced by market women. This will also make tentative suggestions for interventions to address the current situation in closed and open air public markets of Kampala, Uganda.

## METHODOLOGY

### Study Setting/Site

The research study was conducted in Nakasero market, Central Kampala Uganda. Kampala central division is one of the five divisions that make up Kampala the capital of Uganda. The division comprises the central business district of the largest city in Uganda and includes the areas of old Kampala, Nakasero, and Kololo. These areas are the most upscale business and residential neighborhoods in the city. The central division is gifted by a number of public markets including: real best fresh foods (Nakasero), St.balikuddembe(Owino), Usafi, Wandegeya and others. Nakasero market also called real best fresh foods Uganda is a market in Kampala, Uganda, located at the foot of Nakasero hill. It sells fresh food, vegetables and fruits, textiles, shoes and cheap electronics. Nakasero market is located 50 meters off the Entebbe Road(citytours 2019).

### Study population

The study was carried out among female market traders in Nakasero market, central Kampala. These women deal mainly in selling fresh fruits, vegetables and foods in stalls, road-sides and streets of the market.

### Inclusion criteria

Female market traders, who were above the age of 18 years and consented, participated in the research study.

### Exclusion criteria

Female market traders who were too busy and others who rejected to be interviewed during the time of data collection were excluded from the study.

### Study design

The study employed a mixed method approach where both quantitative and qualitative data was collected at the same time. The quantitative part was a cross sectional study while the qualitative was a qualitative descriptive design. Quantitative methods were used to address all the three specific objectives and qualitative for only objective 1: - to examine the occupational hazards experienced by female market traders in Nakasero market that was answered by both qualitative and quantitative methods.

### Sample Size and sampling procedures

For quantitative part, a sample size of 183 female market traders aged over 18 years was estimated using Kish Leslie’s formula for calculating sample sizes (Kish 1959) in cross sectional studies.

The considerations for sample size calculation included anticipated prevalence of obesity as a health problem among market traders from previous studies in public markets in Lagos Nigeria by Odugbemi et.al (obesity (12.3%) (Odugbemi, Onajole, and Osibogun 2012)), 95% confidence limits, 5% precision and non-response rate of 10%.

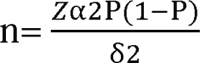

Where n = sample size estimate of female market traders.

Zα= Standard normal deviate at 95% confidence interval corresponding to 1.96

P =anticipated prevalence for obesity as a health problem among market traders from previous studies in public markets in Lagos Nigeria by Odugbemi et.al (obesity (12.3%) (Odugbemi, Onajole, and Osibogun 2012)

δ = Absolute error between the estimated and true prevalence of occupational health hazards among female market traders (5%).

*n* = (1.96)2[0.123(1−0.123] (0.05)2

= 3.8416X0.107871/0.0025

= 166 respondents.

Considering a non-response rate of 10% N= 10/100 X166=17

Therefore; n = 166+17

= 183 sample size

For sample size that participated in qualitative study. Four key informants were interviewed. The key informants were the leaders of female market traders and the market administrators because they were believed to be having vast information about their subordinates.

Simple random sampling was used for the selection of female market traders to participate in quantitative study. Purposive sampling was used to select a number of key informants that participated in qualitative data collection.

### Data collection Procedure

Data were collected between 5^th^ may and 25^th^ July 2022 from those respondents who met eligibility criteria.The languages that were used in both quantitative and qualitative interviews are English and Luganda for effective communication.

For quantitative part of study, the principle researcher collected this data through face to face interviews conducted at the point where female market traders were selling their commodities within the stalls and road-side. Informed consent was obtained from each participant explaining further details about the study and they signed the consent form. A semi-structured questionnaire was used to collect respondent information on: - socio-demographic and economic factors like age, marital status, highest level of education completed, previous occupation, concurrent occupation, main occupation and approximate income per month, information on work-related factors: - working hours, physical activities involved in daily, unsafe practices (non-use of PPE and poor hand washing practices);environmental factors like issue with sanitary facilities, market structures and working space adequacy; information on occupational hazards and health effects faced by female market traders.

Qualitative data collection involved obtaining informed consent first from the key informant before data collection. Four key informant interviews (KIIs) were conducted with the female leaders of various departments in the market probed to provide information using a designed key informant guide. The perspective of the informant with regard to various occupational hazards in the public markets was sought.Verbatim notes and transcript recordings were got from the key informants by recording and writing down notes on key words identified. Qualitative methods were used to supplement quantitative research in some of the specific objectives.

### Data collection Tools

For quantitative part, selected female market traders were interviewed using an interviewer administered semi-structured questionnaire developed from literature on OSH questionnaire (Design and trial of a new questionnaire for occupational health surveys in companies) by A. N. H. Weel and R. J. Fortuin(Weel and Fortuin 1998) and a study on women’s occupational health and safety in the informal economy: Maternal market traders in Accra, Ghana by Joyceline Amoako(Amoako 2019).

For qualitative part, semi-structured key informant interview guide was used for collecting data from key informants. The key informant guides included female market trader’s perspectives on various occupational hazards faced by market women. Both English and Luganda versions of questionnaires were developed and used for effective data collection.

### Quality control Measures

Quality control was ensured by site visit to discuss with authorities, pre-testing and translation of data collection tools, and conducting data checks. The principal researcher was fluent in both English and Luganda languages hence effective data collection. The data collection tools were all translated back and forth into Luganda to ensure uniformity and accuracy of the dialogues.

### Study variables

The dependent variables were health effects faced by female market traders.

Independent variables were the occupational hazards and factors contributing to health effects. Factors that were assessed included mainly the work-related and environmental factors as well as the socio demographic factors. Socio-demographic and economic factors: age, marital status, education level, approximate income per month, main occupation in the market etc. Environmental factors like status of market structures (poor or good housing conditions) and space, sanitary facilities conditions and adequacy, and others. Work related factors such as unsafe practices, and work experience among female market traders were assessed.

### Data management

Raw data and dialogue reports were revisited repetitively to ensure accuracy and quality control. Manual and electronic data storage was performed.

Qualitative data was handled and managed manually and electronically. Interviews were conducted in English and were recorded on Android-equipped smart phones with the help of KII guides.

For quantitative analysis data collection was done by hard copy questionnaires, then data entered and cleaned using EPIDATA version 3.02. It was then transferred to STATA version 14.0 and then Microsoft excels for cleaning and analysing.

### Ethical considerations

Permission to conduct this study was got from Higher Degrees Research and Ethics Committee of Makerere University School of Public Health and Nakasero market administration offices to allow for the research to be conducted at their sites.

Participation was completely voluntary. Written consent was obtained from all study participants after explaining the purpose of the study. During the interview, care was taken not to use negative words and body language which may unintentionally create stigma.

For the KIs, the interviews were conducted in a convenient area of their choice in the offices that allowed privacy.

### Data analysis

Univariate analysis was done for all three objectives. Results were presented in form of pie charts, frequency tables, bar and column graphs. Objective one was analyzed qualitatively using deductive thematic analysis approach.

After data collection from the KI interviews, qualitative analysis was done using deductive thematic analysis with each interview recording being transcribed verbatim by the principle researcher. For convenience to the participant, the data collection was conducted at their place of work at a time that fitted their schedules (when not very busy). A total of 24 codes for KIs were initially generated. Additionally, patterns and themes that resulted from the codes that converged and diverged were recorded. The categorized data was used to identify the main themes of the results. These categories and sub-categories were used to provide information relevant to the study that helped explore and clarify the research questions.

Codes were developed from objective of the study and transcribed data, and then entered into the ATLAS ti version 8 software for analysis. The software developed codes which were reviewed by the researcher and enabled categorization of the study findings. Using deductive thematic analysis, the categorized data was used to develop main themes which made the final results of our study. Finally, the categories were grouped together into overarching themes based on the researcher’s understanding of the data. 8 convergent themes and4 divergent themes emerged from the coding and categorization. A total of 4 key themes emerged from this data analysis. These included; 1 Biological hazards, 2 Ergonomic hazards, 3 Physical hazards, 4 psychosocial hazards. These themes were then documented as qualitative outcomes of our study addressing objective 1: To examine the occupational hazards experienced by female market traders in Nakasero market, and consequent quotes attached to back up our study findings.

Table 1 below shows the coding framework used to arrive at the themes.

**Table 1:** Summary of themes and subthemes.

| Codes generated by key informants | Sub themes that emerged | Top four themes |
| --- | --- | --- |
| KII-24 codes | 08 convergent themes<br>04 divergent themes | 1) Biological hazards<br>2) Ergonomic hazards<br>3) Physical hazards<br>4) Psychosocial hazards |

**Table 2:** a table of frequency and percentages of exposure history to biological hazards among female market traders.

| **occupational hazards variables** | **Freq**<br>**(n=183)** | **P**<br>**ercent** |
| --- | --- | --- |
| respondent ever exposed to occupational hazards | 174 | 95.08 |
| biological hazards |  |  |
| **Exposed to covid-19 virus | 19 | 10.92 |
| **frequent mosquito bites in the market | 152 | 87.36 |
| **UTIs' causing microorganisms from dirty toilets | 140 | 80.46 |
| Others | 63 | 36.21 |
| **multi-response variables |  |  |

**Table 3:** respondents who have experienced work-related stress in the market.

| Respondent experienced any work-related stress | frequency<br>(n=183) | percent<br>age(%) |
| --- | --- | --- |
| yes | 180 | 98.36 |
| no | 3 | 1.64 |

## RESULTS

This chapter presents the quantitative and qualitative study findings of an assessment of the occupational hazards and factors contributing to the health effects faced by 183 female market traders in Nakasero market, central Kampala Uganda. It starts with findings on occupational hazards experienced by female market traders; followed by findings on health effects faced by female traders arising from exposure to hazards in the market and finally factors contributing to the occupational health effects faced by female traders.

### OCCUPATIONAL HAZARDS EXPERIENCED BY FEMALE MARKET TRADERS

#### Biological hazards

As presented by the table 4 below, majority of respondents174/183 (95%) reported that they had previous exposure to various hazards in the market. The table indicates that majority reported of being exposed to frequent mosquito bites in the market (87.36%) and UTIs’ causing micro-organisms from dirty toilets (80.46%).

**Table 4:** physical activities involved in by the respondents in the market.

| physical activity | Fre<br>quency<br>(n=183) | Per<br>centage<br>(%) |
| --- | --- | --- |
| involved in sorting items daily | 130 | 71 |
| involved in packing daily | 175 | 95.6 |
| Involved in lifting items | 135 | 73.77 |
| involved in other activities | 69 | 37.7 |
| *multi-response variables. |  |  |

### Theme I: biological hazards

The various biological hazards experienced in Nakasero market were reflected in qualitative comments of the key informants. From carrot selling department, a head of department and female trader in Nakasero market shared her thought on biological hazards faced in by market women in this area.

> “I beg the ministry of health to give us free mosquito nets. At home i sleep in mine but while in the market overnight i really suffer from mosquito bites. I still get malaria from time to time even my fellow female traders here in this market are always sick with malaria fever. During covid-19 lockdown it was even worse; we used to sleep in the market every day so we really faced it rough with mosquito bites at night. Another issue is market toilets, I’m shy to talk about this since you are a man but hope you understand. Many market women complain of itches in the private parts after using the toilets, even me myself am a victim. More toilets are needed but more so the cleanliness should be a priority” (Head of department for carrot sellers, 4 years in service).

### Ergonomic hazards

Figure 1 below highlights the ergonomic hazards that cause musculoskeletal problems, injuries and pain. It indicates that majority of the respondents’ sit/stand for so long in awkward postures 89.07% (163/183) and experience repetitive movements (59%), lifting items and over-bending that exacerbates musculoskeletal effects.

**Figure 1:**
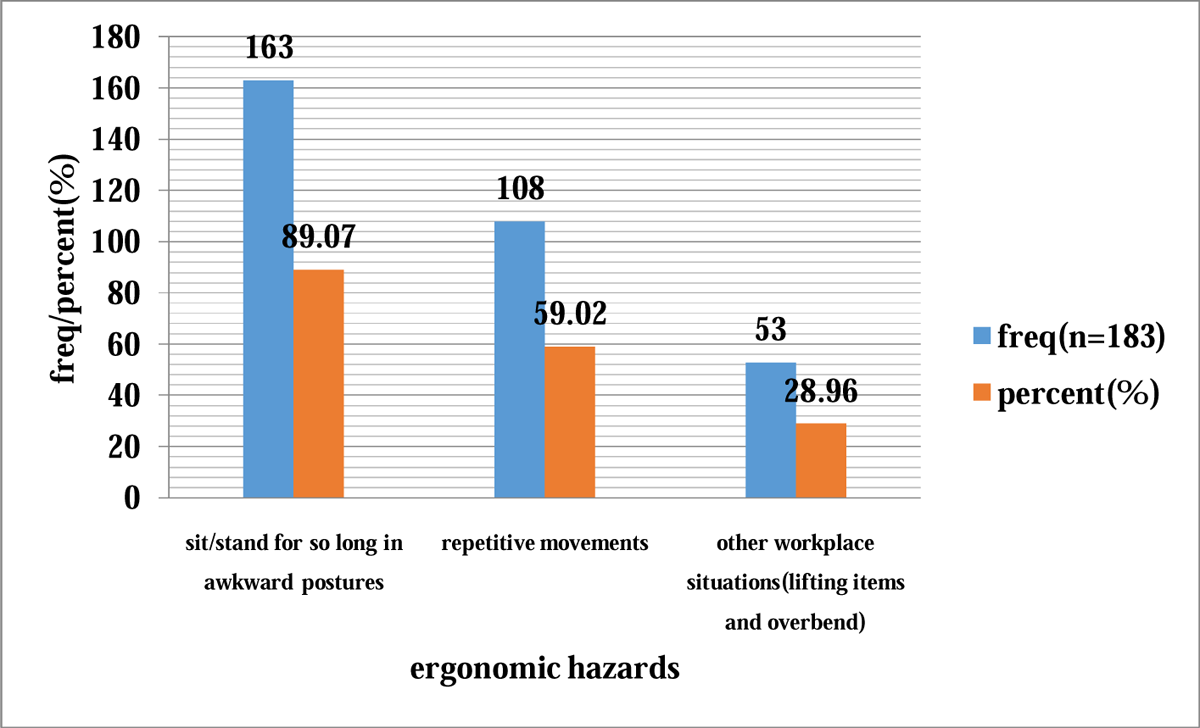
a column graph showing the respondents’ experience on ergonomic hazards that predisposes them to musculoskeletal injuries.

### Theme II: Ergonomic hazards

One of the most mentioned work hazards among key informants was the ergonomic hazards suffered from by market women. The informants talked about their working conditions which usually resulted in fatigue, body pain and other health issues. A female market trader and leader of banana sellers department talked about the strenuous activities female market traders’ face.

> “The work that market women do here is very strenuous. Waking up early or sleeping over in the market, carrying luggage .e.g. banana baskets, sorting, arranging and packing items on sale. We really move a lot, this way that way aah.…! It’s just tiring. One has to bend to pick up for buyers and this affects our waist badly. You have to bend more than ten times when dealing with difficult buyers because they reject the selection you make for them. Get tired by noon but you still sell until everything has been bought.” Leader of banana sellers department, 9 years in service said.

### PHYSICAL HAZARDS

As depicted on figure 2 below the respondents agreed that they do suffer mostly from physical hazards like excess noise from people and machines (96.17%) and extreme temperatures from sunshine and heavy rain (87.98%) due to temporary market structures in Nakasero open-air market

**Figure 2:**
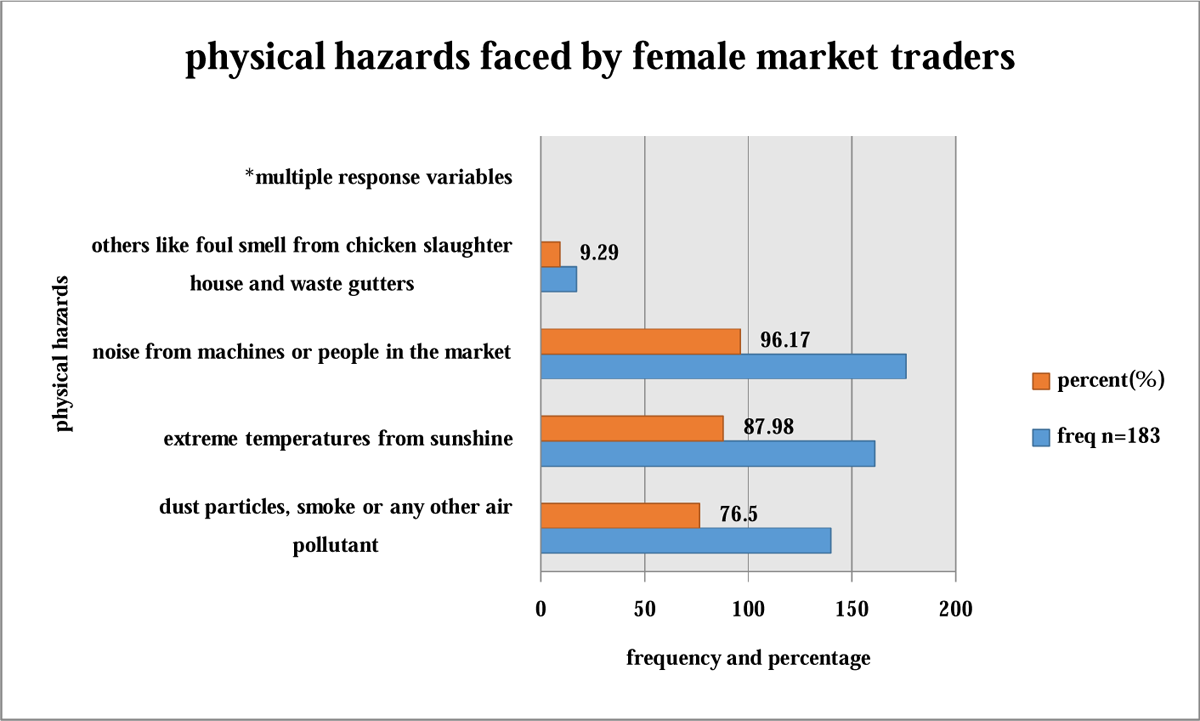
a bar graph showing frequencies and percentages of physical hazards in Nakasero open-air market.

### Theme III: Physical hazards

One of the informants had something to say about the physical hazards and this included: hot temperatures, heavy rains, excess noise and foul smell from chicken slaughter house and waste channels in the market. This was from a head of onion selling department and below is how she narrated to the researcher.

> “We face big trouble during hot days and when there comes heavy rains in this open air market since our stalls are housed by temporary structures like umbrella. For us women we sweat a lot when we feel heat especially during a sunny day at noon. Too much noise in the market is yet another problem all market traders will tell you about. Not forgetting the bad smell we get chicken slaughter house and waste trenches, hmmmm…….! It’s just unbearable.” Head of onion sellers department, 3 years in service said.

### Psychosocial hazards among female market traders

Almost all female traders 98% (180/183) in general reported experiencing work-related stress while in the market due to various causes as revealed by table 5 below.

**Table 5:**
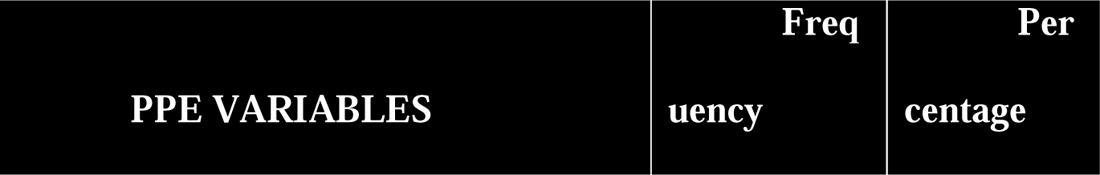
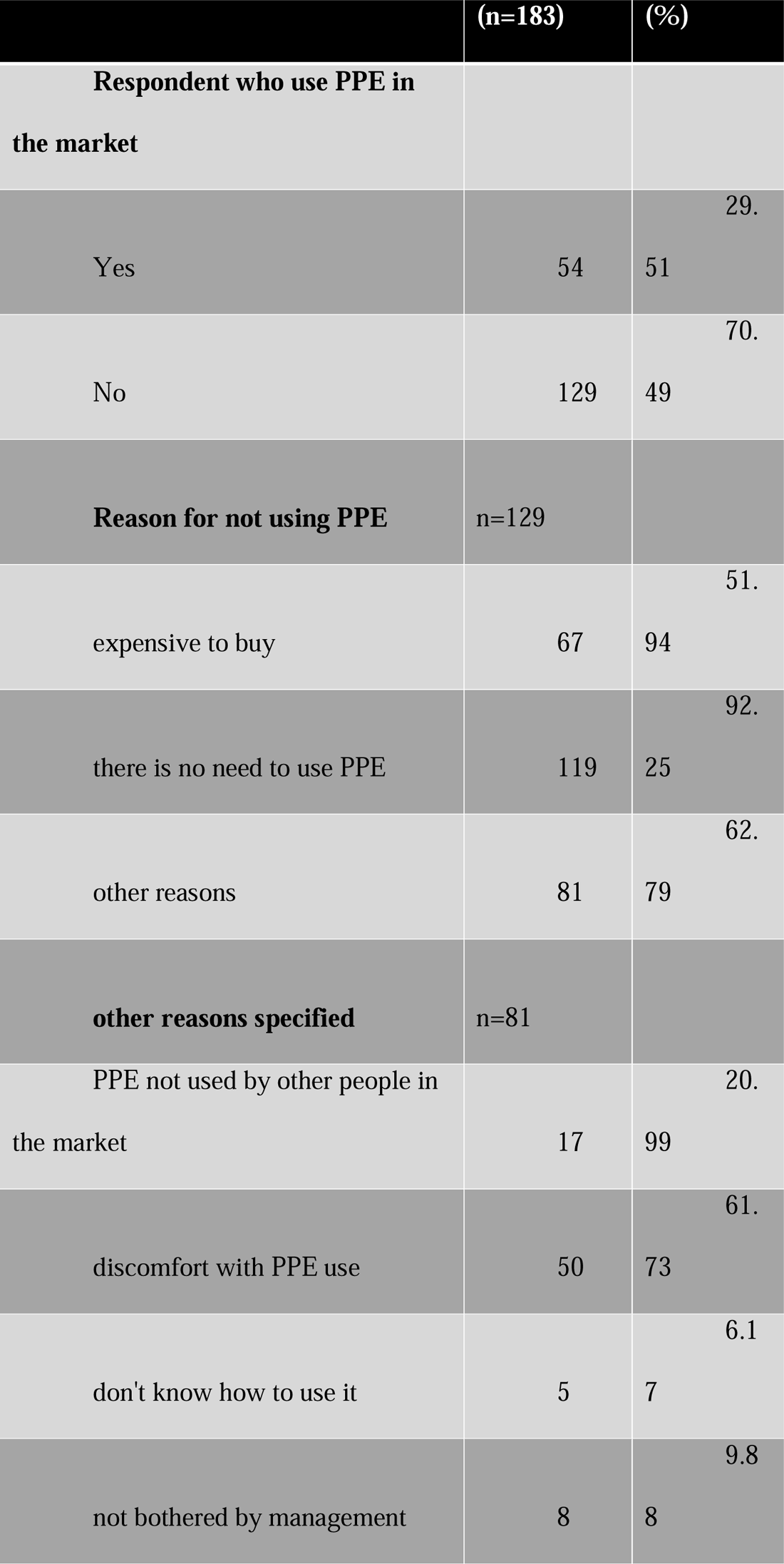
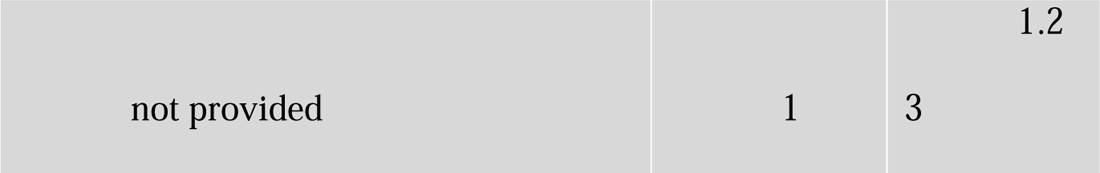
Frequency and percentage distribution of PPE non-use related factors among female market traders.

### Causes of stress among female market traders

The information provided in the figure 3 below, indicates that the main causes of stress among female market traders were working with clients with challenging behaviors 95% (171/183), long working hours without rest/sleep 87.78% (158/183). Incurring business losses and harsh treatment by KCCA and market authorities was yet frequently reported by many respondents as a cause unending stress

**Figure 3:**
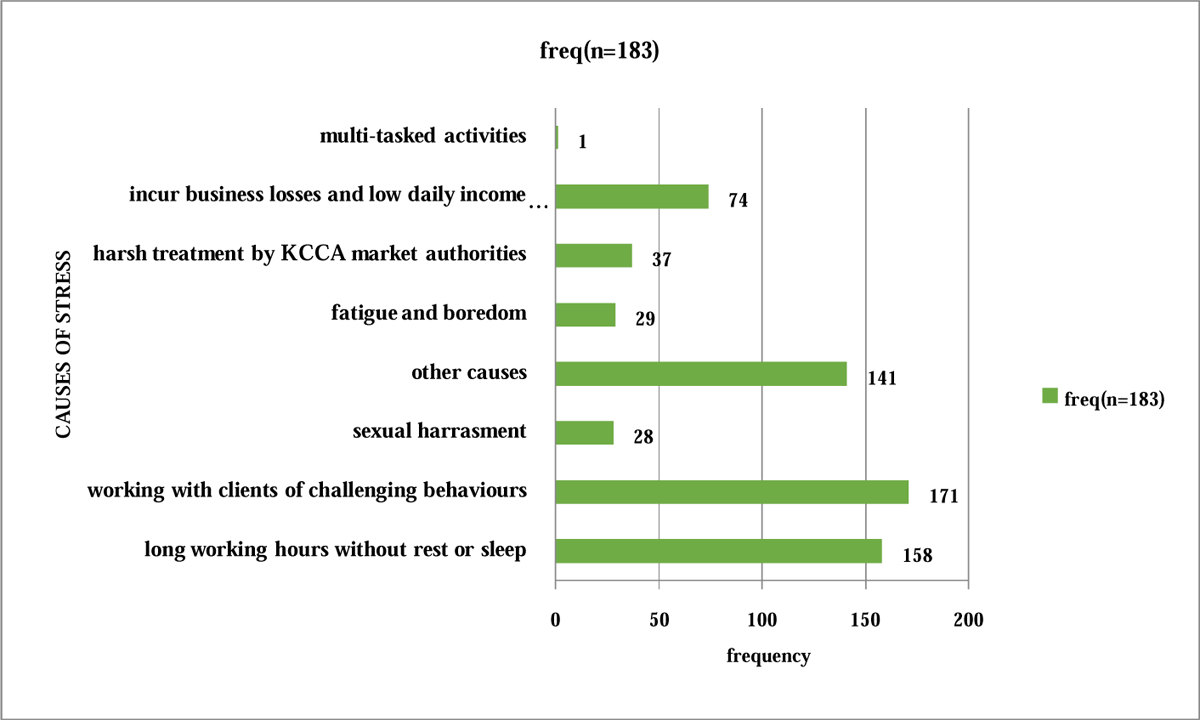
A bar graph showing causes of stress among female market traders.

### Theme IV: psychosocial hazards

Almost all of the key informants (3/4) interviewed mentioned the issue of psychosocial and emotional stress among female market traders in the market. Some believed that the crisis in the national economy trickled down to affect their businesses by first, raising the prices of goods and second, reducing the purchasing power of their customers hence low daily earnings. This imbibed a state of fear, worry and stress on these market women. Below was the quote from one of the key informants (overall chairperson for market women) who reported that female market traders are faced by stress, worry and financial problems.

> “Working in the market is very exhausting. We are always worried and stressed with this market business. At times you earn less and make more losses. One also gets frequent headaches and sleeplessness. May be we overwork or have no time enjoy life. The economy is also hard; I haven’t even sold anything since morning.” Says the chairperson for market women, 7 years in service.

### HEALTH EFFECTS FACED BY FEMALE TRADERS ARISING FROM EXPOSURE TO HAZARDS IN THE MARKET

As illustrated in the figure 4 below majority of the respondents agreed that the main health effects they suffer from in the market were musculoskeletal pain and injuries (92.35%) followed by respiratory problems (63.93%). Over half of the respondents (52.46%) reported to have frequently suffered from malaria illnesses, UTIs (28.42%) and 22.4% reported of past history with Covid-19. Other health effects reported were headache, lower abdominal problems and mental illness though undisclosed.

**Figure 4:**
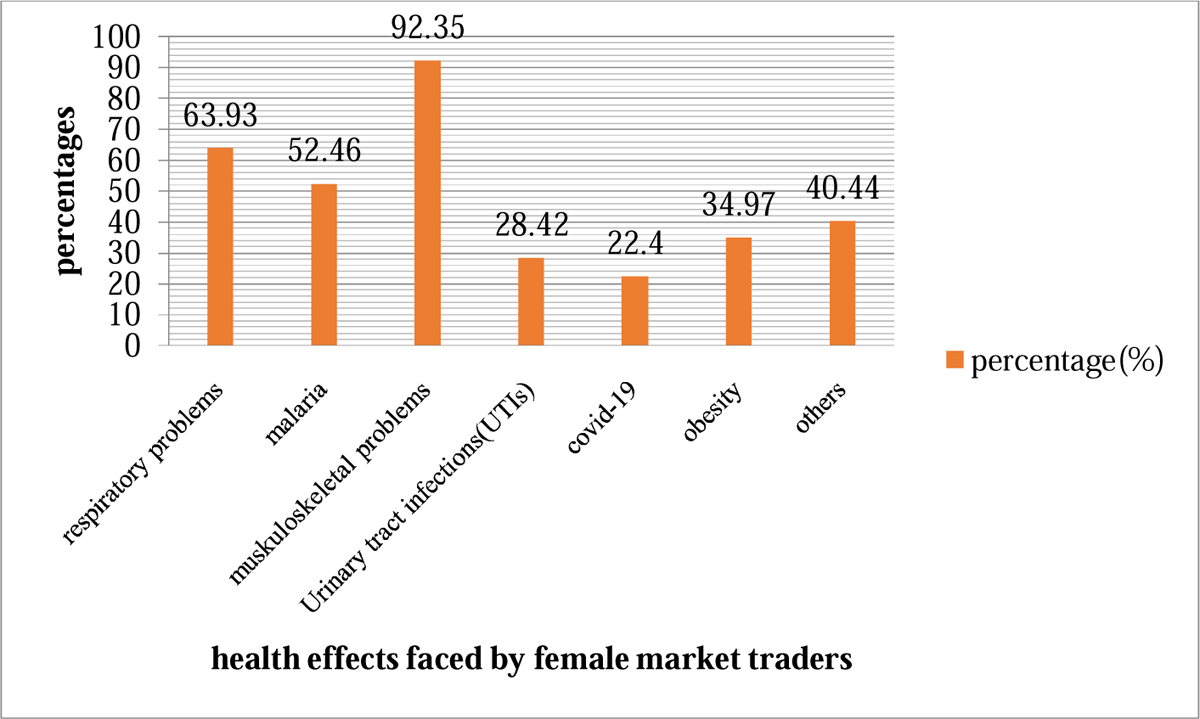
a column graph showing the various health effects reported among female market traders in Nakasero market.

### FACTORS CONTRIBUTING TO THE OCCUPATIONAL HEALTH EFFECTS FACED BY FEMALE TRADERS

#### Work related factors

##### Working hours spent in the market per day

Figure 5 below shows that almost half (86/183) 47% of the female market traders spent more than 12 hours in the market, followed by (76/183) 41.5% respondents that spent 8-12 hours in the market. Among those who reported of frequent musculoskeletal pain and injuries, they spent long working hours in the market per day. Ergonomics and stress were deemed dependent on working overtime in the market as revealed in previous studies and as in the next section (discussion).

**Figure 5:**
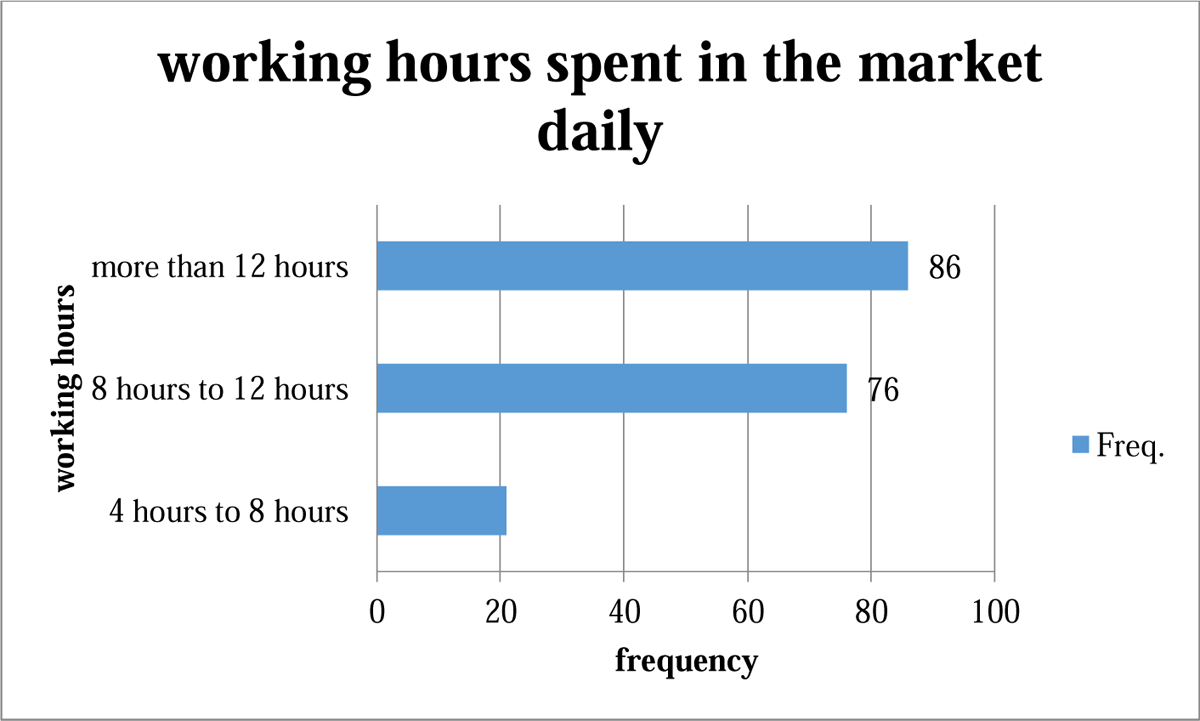
a bar graph showing the working hours spent by the respondents in the market per day.

**Figure 6:**
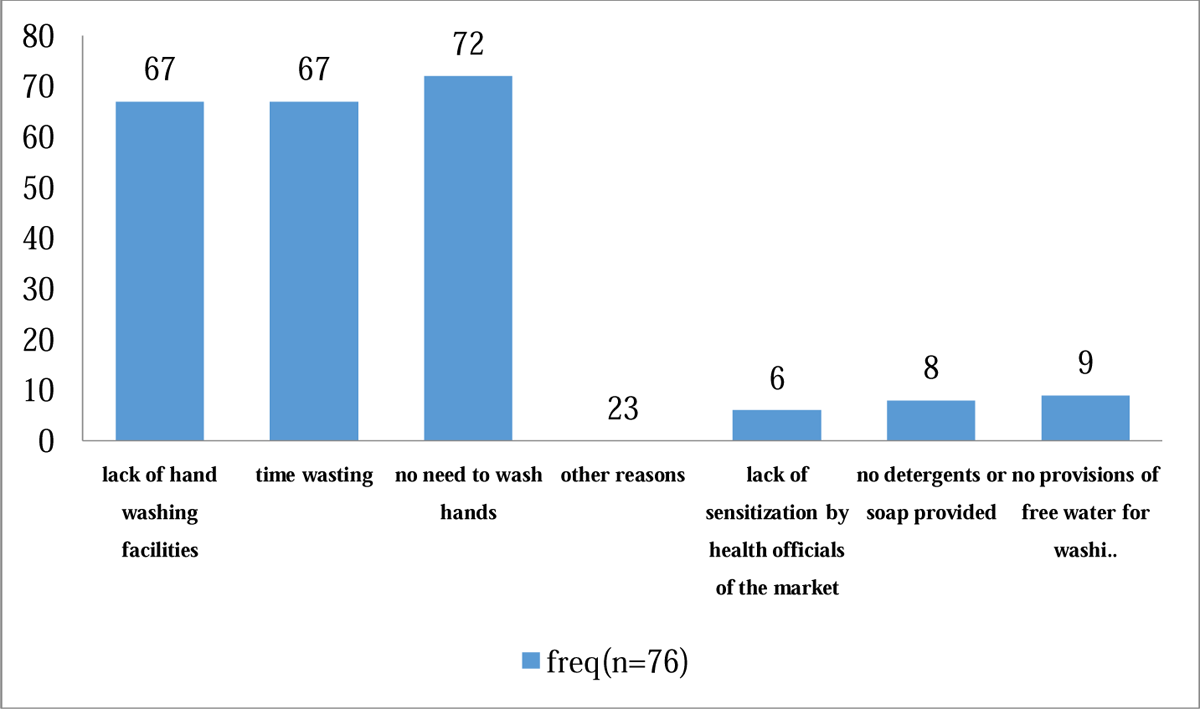
a column graph showing the respondent’s reasons for not washing hands in the market.

### Physical activities involved in daily by the respondents in the market

Table 4 below show that majority 71% (130/183) of the female market traders were actively involved in sorting items on sale daily, 95.6%(175/183) reported that they do packing of commodities repetitively and that was tiresome, 73.77%(135/183) lifting items sold to customers to their vehicles in the park yard, only 37.7% of the respondents revealed that they are involved in other activities like peeling, washing fruits and food delivery. These physical activities were reported to be contributing a lot the musculoskeletal pain and injuries faced by female market traders in the long run while in the market.

### Unsafe practices

#### PPE NON-USE AMONG FEMALE MARKET TRADERS

Table 5 below shows that70.5% of respondents were not using PPE in the market, majority 92.25 %(119/183) of the respondents not using PPE in the market believed that there is no need to use PPE whereas other reason that dominated the non-users of PPE was discomfort 61.73%(50/81). PPE non-use is a key contributory factor for the spread of communicable diseases thus explaining the high prevalence’s of respiratory health problems (63.93%) and covid-19 (22.4%) as reported by female market traders.

### Poor hand-washing practices among female market traders in the market

Table 6 below shows that 42% (76/183) of female traders in Nakasero market were not washing hands while in the market. Lower abdominal problems and diarhea though less frequently mentioned as others in health effects faced by these market women were arising as a result of this poor hand washing practice.

**Table 6:** hand washing practice among female market traders.

| Respondent who were washing hands in the market | frequency(n=183) | Percentage (%) |
| --- | --- | --- |
| yes | 107 | 58.47 |
| no | 76 | 41.53 |

**Table 7:** Respondents who faced difficulties with usage of sanitary facilities in the market.

| Respondents who faced challenges<br>with market sanitary facilities | frequency<br>(n=183) | percent<br>age(%) |
| --- | --- | --- |
| yes | 174 | 95 |
| no | 09 | 5 |

The findings on the reasons for not washing hands in the market as reported by some female market traders revealed that majority,88.16%(67/76) reported that lack of hand washing facilities in the market and perceived time wasting were the key reasons for not washing hands. 94.74 %(72/76) were not practicing hand washing in the market saying that there is no need to do so.

### Environmental factors

#### Challenges with usage of sanitary facilities in the market

Table below shows that 95% (174/183) of the female market traders reported that they were facing challenges in using the sanitary facilities in the market. This predicted a high risk of exposure to occupational biological hazards for instance UTIs’ causing microorganisms from unsanitary toilets.

Figure 7 below reveals that majority 91.95 %(160/174) of the female market traders have frequently suffered from long time taken awaiting access to sanitary facilities in the market. Dirty toilets/washrooms 89.08 %(155/174) was yet another problem that was common in the markets as reported by respondents. Insufficient water on some occasions was also mentioned by many respondents 71.3% (124/174) as one of the major challenges affecting the use of sanitary facilities in the market

**Figure 7:**
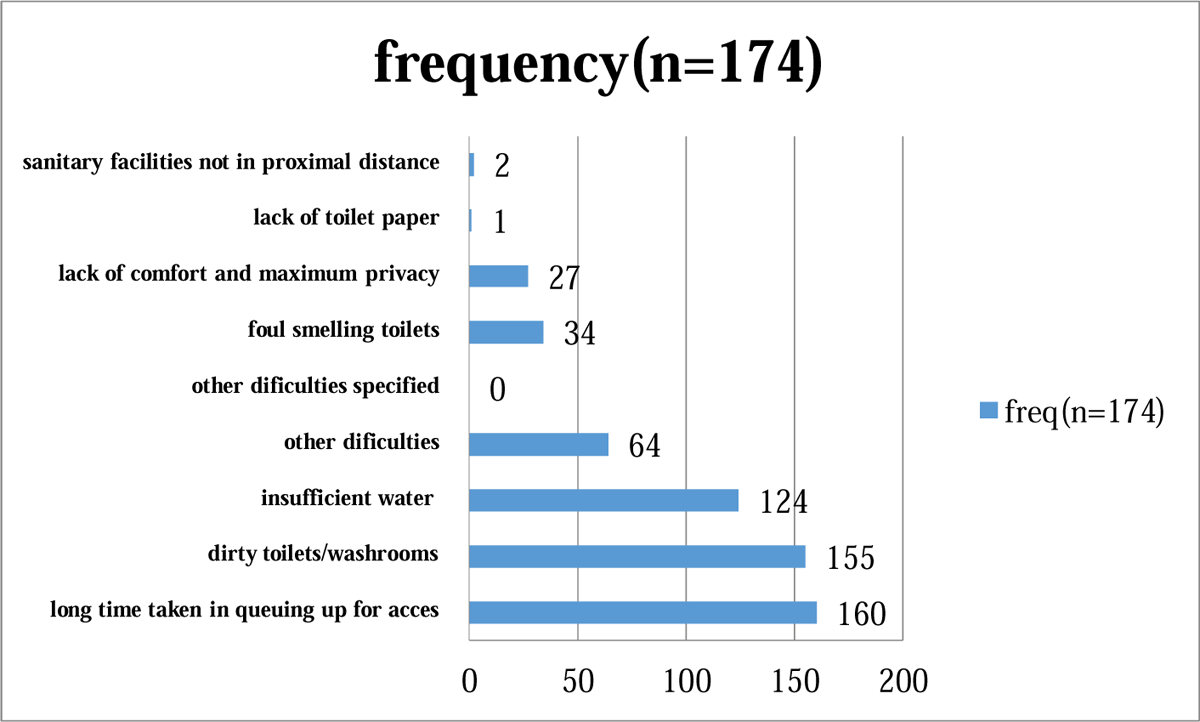
a bar graph showing the challenges suffered from by respondents in using market sanitary facilities.

### Evaluation of status of market structures

Figure 8 below shows that more than half of the female market traders 51.37% (94/183) reported the market structures inform of stalls, housing materials and floors are fair and as such needs improvement. It was necessary to understand the market structures’ status because bad shelter exposes the traders to environmental hazards for this case extreme weather conditions such as hot sunshine and heavy rains.

**Figure 8:**
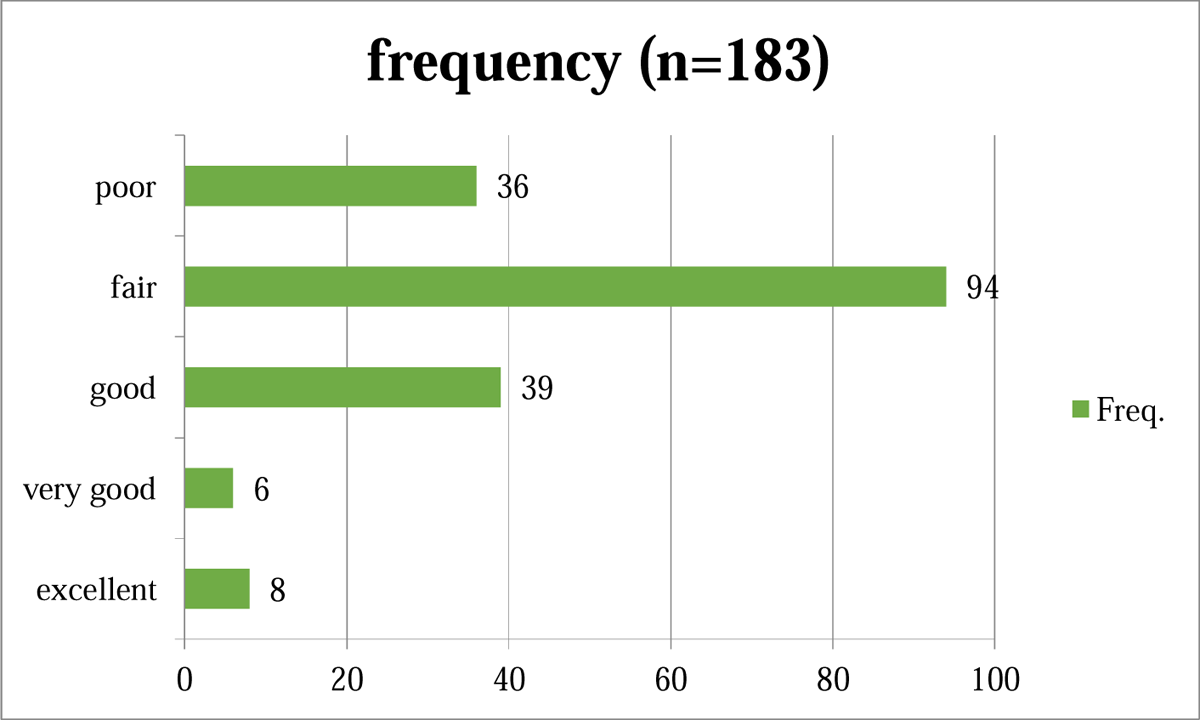
a bar graph showing the evaluation of status of market structures by the respondents.

Figure 9 below shows that majority 77% (138/183) of the respondents cried of working space inadequacy in the market hence overcrowding was the implication of this.

**Figure 9:**
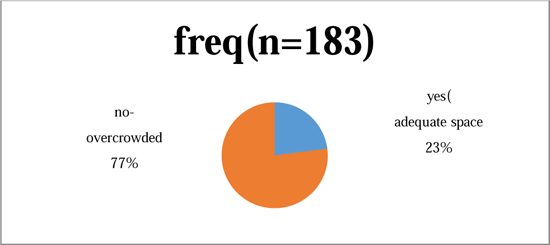
a pie chart showing response on working space adequacy/overcrowdings among female market traders.

## DISCUSSION

### OCCUPATIONAL HAZARDS IN MARKET FACED BY FEMALE MARKET TRADERS

Respondents reported that they do suffer mostly from physical hazards like excess noise from people and machines (96.17%) and extreme temperatures from sunshine and heavy rain (87.98%) due to temporary market structures in Nakasero open-air market. This study was similar but with higher cases reported compared to a research by Pick et al. (2002) on the health hazards encountered by female market and street vendors in Johannesburg, South Africa. Pick et al found out that more than half (52%) of the women reported that they were not happy with the working environment for reasons extending from lack of shelter, noise from many people in the market (26%) and having to clean the area themselves (24%)(Pick, Ross, and Dada 2002). On assessment of biological hazards, frequent mosquito bites in the market (87.36%) and exposure to UTIs’ causing microorganisms (80.46%) were the major biological hazards reported by female market traders in the market. Only a minority number (10.92%) reported of history of exposure to Covid-19 virus in market.

The study findings on the ergonomic hazards indicated that majority of the market women sit/stand for so long in awkward postures 89.07% (163/183, experience repetitive movements(59%); lifting items and over-bending that exacerbate musculoskeletal illnesses, injuries and pain. This finding agreed with study done by Avotri and Walters among market women. They discovered that enormous amount of work, bad postures and financial insecurity were found to contribute to the women’s anxiety, which in turn was linked to the tiredness, lack of sleep and bodily aches and pain that many of them experienced as chronic health difficulties (Avotri and Walters 1999). To reduce on the risk of ergonomic hazards among the female market traders’ therefore occupational health education and sensitization should be intensified to improve on awareness and practice of safe workplace procedures and conditions.

Almost all female traders 98% (180/183) in general reported experiencing psychological stress while in the market due to various causes. The main causes of stress among female market traders as depicted from the findings were working with clients with challenging behaviours 95% (171/183) and long working hours without rest/sleep 87.78% (158/183). Incurring business losses and harsh treatment by KCCA and market authorities was yet frequently reported by many respondents as a cause unending stress. This study correlates with a study done by Alfers and Rogan in 2015 that found out that stress among female market traders was due to long working hours and the lack of labour protection. This meant that they are not able to care for themselves adequately either; for example, by taking time off work to visit the doctor when they are ill(Alfers and Rogan 2015). This therefore necessitates the need for the government to intervene by establishing legal protections on these female traders in the market as regarding taxation, working time and working conditions.

A research study done by JoselineAmoako (2019) on occupational health hazards faced by maternal market traders in Accra, Ghana formulated a research question that asked about the health hazards that exists in market places and nearby streets. The participants mentioned a lot but the hazards mentioned more frequently in their responses were poor sanitation, fire outbreaks, overcrowded places, harassment from local officials, extreme weather conditions and excessive workloads(Amoako 2019). This study was closely related to my study but had no statistical findings since it was only qualitative and yet again that study focused on only maternal market traders yet all women are the vulnerable to health hazards and need special attention.

### HEALTH EFFECTS FACED BY FEMALE MARKET TRADERS

Research findings on health effects among female market traders depicted that the main health effects suffered from in the market were musculoskeletal pain and injuries(92.35%) followed by respiratory problems(63.93%). A descriptive cross-sectional survey conducted among all the female traders in Sango market, Ibadan in, April 2003 by Balogun.et.al identified the common health problems of women traders in Sango market Nigeria and their work conditions. The most commonly reported health problems were muscular and joint pains by 37.4%, 33.8% had symptoms suggestive of malaria and 23.5% had chronic low back pain(Balogun and Owoaje 2007). This shown a lower prevalence of musculoskeletal pain 37.4% in Nigerian market as compared to a Ugandan market 92.35%. This could imply that market women in Uganda are more exposed to occupational factors like immerse workload, bad postures and long working hours.

### FACTORS CONTRIBUTING TO THE OCCUPATIONAL HEALTH EFFECTS FACED BY FEMALE MARKET TRADERS

Results on work related factors depicted that majority (86/183) 47% of the female market traders spent more than 12 hours in the market. Musculoskeletal pain injuries and stress were frequently reported among those who worked for longer hours as from previous studies. Long working hours without rest or sleep have been earlier on discovered in a similar study done by(Balogun and Owoaje 2007) as a factor contributing to muscular and joint pain and injuries. This study by Balogun and Owoaje indicated that musculoskeletal problems were highest among those who spent eight to ten hours per day in the market (20%). A study by Alfers and Rogan described long working hours as a double burden to market women and likely due to lack of or few labour protections(Alfers and Rogan 2015).Long hours at work may extend the time into the evening and night when they must look after the needs of their own household.

The research findings on unsafe practices among the female traders in Nakasero market revealed that majority of the respondents were not using PPE (70.49%) and minor though a reasonable number (42%) reported of not washing hands in the market. This could expose the female traders to workplace risks of pathogens that cause diarrheal diseases like cholera, dysentery etc, spread respiratory infections like corona virus, cold flu, TB and many more threats. These findings of the safety practices could therefore imply that the less preparedness among the market women could pose a risk of outbreaks of infectious diseases that can easily spread through poor hygiene and direct contacts. Respiratory problems could be prevailing in markets due to overcrowdings, poor sanitation and lack of proper use of PPE.

On evaluation of the environmental factors in Nakasero market, the study found out that the market women were facing problems with use of sanitary facilities on issues like few stances, dirty toilets and water insufficiency at times. This could imply that female traders are at increased risk of urinary tract infections (since they are the vulnerable population.Women are more susceptible to genital ulcerative diseases (GUDs) and sexually transmitted infections (STIs) than men due to “hormonal changes, vaginal microbial ecology and physiology” (Quinn & Overbaugh, 2005). I also found out that majority of the participants 51.37% reported the market structures were fair, still lacking and many cried of space inadequacy. This is similar to a study by (Pick, Ross, and Dada 2002) that shown that more than half (52%) of the women reported being unhappy with the working environment for reasons extending from lack of shelter and dirt (34%).The study revealed that majority 77% (138/183) of the respondents cried of working space inadequacy in the market hence overcrowding was the implication of this.

## CONCLUSION

Female market traders continue to face several occupational hazards and health effects in public markets within Uganda. Mainly biological hazards such as mosquito bites in the market and exposure to UTIs causing micro-organisms from dirty toilets, ergonomic hazards such as sit/stand for so long in awkward postures, repetitive movements; lifting heavy items and over-bending that exacerbate musculoskeletal illnesses, injuries and pain. Psychosocial hazards faced by this market women included working with clients with challenging behaviors, long working hours without rest/sleep, Incurring business losses and harsh treatment by KCCA and market authorities. The major health effects identified by this research were musculoskeletal pain, malaria, UTIs, headache and lower abdominal problems. The factors responsible for experiencing hazards include not wearing all necessary protective equipment; not washing hands in the market, working overtime, sit/stand for long in bad postures, problems with use of sanitary facilities on issues like few stances, dirty toilets and water insufficiency at times, poor market structures and working space inadequacy hence overcrowdings in the market.

### RECOMMENDATIONS

The government should work together with market authority (KCCA) to install and ensure clean sanitary facilities, improved market structures, hand washing facilities and many more basing on the assessment made in regard to occupational hazards.

The Ministry of Gender, Labour and Social Development should focus more on social and labour protections of these vulnerable population (market women) by enforcing employment protection regulations where necessary and introducing new legislation and standards to protect the health of working women in these informal sector.

The government should necessitate provision of health centers near markets: Available and accessible high quality health-care services, affordable to all, are vital determinants of female trader’s health.

Literacy and education on occupational health and safety: female market traders should be health educated on potential risks and hazards in the public markets that may pose a threat to their health.

Market authority and management need to provide enough and subsidized stalls to help solving the problem of working space inadequacy and overcrowding.

The government should dive in promoting empowerment and training of safer practices and behavior of these female market traders through local leaders and health servants within market jurisdiction.

This informal economy needs the Ministry of Gender, Labor and Social Development to assist with social and labor protection of these female market traders by funding and educating on market business. Government and the market authorities (KCCA) should therefore consider creating more market space for instance using multi-storey buildings.

## Data Availability

All data produced in the present work are contained in the manuscript

## ACKNOWLEDGEMENT

I am deeply grateful to the MAKERERE University and the government of Uganda for according me a special opportunity to pursue a Bachelor’s degree in environmental health science at the school of public health on full Scholarship. My Heartfelt gratitude goes to my supervisor Dr. Marianna Agaba Nyangire for kindly and patiently guiding me throughout the process of my research and, for also mentoring me as regarding further studies in public health and occupational health. With profound gratitude, I acknowledge the dedicated efforts from all my friends that have given me the encouragement and motivation during this research, I appreciate you all. To all the lecturers that imparted knowledge to us, I am immensely grateful for the opportunity to learn and interact with you all.

My appreciation also extends to the entire 2019 Cohort of BEHS for all the beautiful memories we shared together. I would love to extend my heartfelt gratitude to my family, especially my mother NAMBUBA Beatrice, father late WAMANGA Charles, my wife Joan NAMONO plus my stubborn boy WAMANGA Edgar who encouraged and believed in me during the entire three years of study. Finally, I am most grateful to God almighty for the strength, favor, grace, wisdom, good health and provision for the entire three years, Honor and glory back to you

## ACRONYMS AND ABBREVIATIONS

WIEGO: Women In Informal Employment: Globalizing and Organizing

ILO: International Labour Organization

COSDOH: Commission on Social Determinants of Health

KCCA: Kampala Capital City Authority

WHO: World Health Organization

PPE: Personal Protective Equipment

MSDs: Musculoskeletal Disorders

OSH: Occupational Safety and Health

LMIS: Labor Market Information Status

SANITATION: the practice of keeping places clean and healthy

SSA: Sub Saharan Africa

SD: standard deviation

KI: key informant

KII: key informant interview

UTI: urinary tract infections.

## References

Adjokatse, IT, T Oduro-Okyireh, and M Manford. 2022. ‘Health Problems Associated with Market Women in Closed and Open Space Market Areas’, African Journal of Applied Research, 8: 97–107.

akinamamawafrica. 2020. ’THE INFORMAL ECONOMY IN THE GRIP OF THE PANDEMIC’. https://www.akinamamawaafrika.org/the-informal-economy-in-the-grip-of-the-pandemic-sharing-experiences-and-strategies-for-a-resilient-recovery/.

Alfers, L., and M. Rogan. 2015. ‘Health risks and informal employment in South Africa: does formality protect health?’, Int J Occup Environ Health, 21: 207–15.

Alfers, Laura. 2009. ’Occupational health & safety for market and street traders in Accra and Takoradi, Ghana’, Women in Formal Employment; Globalising and Organizing.

Amoako, Joyceline. 2019. ’Women’s occupational health and safety in the informal economy: Maternal market traders in Accra, Ghana’.

Avotri, Joyce Yaa, and Vivienne Walters. 1999. “You just look at our work and see if you have any freedom on earth”: Ghanaian women’s accounts of their work and their health’, Social Science & Medicine, 48: 1123–33.

Balikowa, David Ouma. 2020. ’Local markets in Kampala(Advocacy for public space)’. https://healthbridge.ca/dist/library/Uganda_Market_Report_Jan18_compressed.pdf.

Balogun, M. O., and E. T. Owoaje. 2007. ‘Work conditions and health problems of female traders in Ibadan, Nigeria’, African journal of medicine and medical sciences, 36: 57–63.

Bonnet, Florence, Joann Vanek, and Martha Chen. 2019. ’Women and men in the informal economy: A statistical brief’, *International Labour Office*, Geneva.

Chen, Martha Alter. 2001. ‘Women and informality: A global picture, the global movement’, Sais Review, 21: 71–82.

citytours, kampala. 2019. ‘biggest local markets in kampala uganda’.

Flaspöler, E, A Hauke, D Koppisch, D Reinert, T Koukoulaki, G Vilkevicius, L Žemės, M Águila Martínez-Casariego, M Baquero Martínez, and L González Lozar. 2013. ’New risks and trends in the safety and health of women at work’. Waneloba Ronald / Assessing Occupational Hazards and Factors Contributing to the Health Effects Faced By Female Market Traders in Nakasero Market, Central Kampala Uganda / 34

Forastieri, Valentina. 1999. ’Improvement of working conditions and environment in the informal sector through safety and health measures’, Geneva: International Labour Office: 1–17.

Fred, Alinda, Alex Nduhura, and Mukasa Ronald. ’PROMOTING ENTREPRENEURSHIP THROUGH THE MIND-SET CHANGE APPROACH: EMPIRICAL EVIDENCE FROM WOMEN ENTREPRENEURS IN UGANDA’.

Health(COSDOH), Commission on Social Determinants of. 2008. “Closing the gap in a generation: health equity through action on the social determinants of health: final report of the commission on social determinants of health.” In. Geneva: World Health Organization.

Hill, Allan G, Rudolph Darko, J Seffah, Richard MK Adanu, John K Anarfi, and Rosemary B Duda. 2007. ‘Health of urban Ghanaian women as identified by the Women’s Health Study of Accra’, International Journal of Gynecology & Obstetrics, 99: 150–56.

ILO. 2018. ’International Labour Organization. Women and Men in the Informal Economy: A Statistical Picture, 3rd ed.; International Labour Organization: Geneva, Switzerland, 2018.’.

Kevin, Nwanna Uchechukwu. 2019. ‘The Occurrence of Workplace Hazards among Selected Workers in the Informal Sector Kampala Uganda’, Occupational Diseases and Environmental Medicine, 7: 164–75.

Law, Mary, Sandy Steinwender, and Leanne Leclair. 1998. ‘Occupation, health and well-being’, Canadian Journal of Occupational Therapy, 65: 81–91.

Loewenson, René. 2002. ’Occupational hazards in the informal sector—a global perspective.’ in, Health effects of the new labour market (Springer).

Megersa, Kelbesa. 2020. ‘The Informal Sector and COVID-19’.

Odugbemi, T. O., A. T. Onajole, and A. O. Osibogun. 2012. ‘Prevalence of cardiovascular risk factors amongst traders in an urban market in Lagos, Nigeria’, Niger Postgrad Med J, 19: 1–6.

Onkarnath, Chattopadhyay and. 2005. ‘Safety and Health of Urban Informal Sector Workers’, Indian Journal of Community Medicine, 30.

OSINDE, Anthony. 2020. ’Influence of Social Factors on Energy Recovery Options from Solid Waste Generated in Markets: A Case of Central Division of Kampala’, PAUWES.

Pick, W. M., M. H. Ross, and Y. Dada. 2002. ‘The reproductive and occupational health of women street vendors in Johannesburg, South Africa’, Soc Sci Med, 54: 193–204. Waneloba Ronald / Assessing Occupational Hazards and Factors Contributing to the Health Effects Faced By Female Market Traders in Nakasero Market, Central Kampala Uganda / 35

Rajen N Naidoo, Florian M Kessy, louis Mlingi. 2009. ’Occupational health and safety in the informal sector in Southern Africa - the WAHSA project in Tan Rajen N Naidoo, Florian M Kessy, Louis Mlingi Download’.

UBOS. 2020. ’uganda bureau of statistics report’.

Uko, FE, MBUOTIDEM JOHN Akpanoyoro, and JOHN POLYCARP Ekpe. 2020. ‘AN EVALUATION OF THE CONTRIBUTION OF THE INFORMAL SECTOR IN EMPLOYMENT AND INCOME GENERATION IN NIGERIA: THEMATIC APPROACH’, Management, 1.

Weel, ANH, and RJ Fortuin. 1998. ‘Design and trial of a new questionnaire for occupational health surveys in companies’, Occupational Medicine, 48: 511–18.

WHO. 2006. Gender equality, work and health: a review of the evidence (World Health Organization).

worldatlas. 2019. ‘contries with the highest informal economy’.

